# Team-Based Learning Versus Lecture-Based Instruction for Chest Radiograph Interpretation in Physician Associate Education: A Quasi-Experimental Study

**DOI:** 10.64898/2026.02.20.26346418

**Authors:** Kelsey F. Kehrli, Kelly R. Conner, Lauren Eyadiel, Caroline B. Sisson, Natalie Smith

## Abstract

**Background:** Chest radiograph interpretation is a foundational skill in physician associate (PA) education, and competence in diagnostic imaging is an accreditation standard. While a larger body of research on radiology education exists in undergraduate medical education, considerable variability in instructional approaches limits clear conclusions regarding the most effective method. Growing evidence supports the use of active learning strategies in radiology instruction. However, little published research specifically addresses radiology education within PA programs. Team-Based Learning (TBL), an active learning approach grounded in social constructivism that emphasizes preparation, collaboration, and application, may be well suited to teaching image interpretation. This study evaluates the effectiveness of TBL compared with traditional lecture-based instruction for chest radiograph interpretation.

**Methods:** A mixed-methods, quasi-experimental cohort comparison using a pre–post design was conducted with two consecutive PA student cohorts at a single institution. One cohort received a 90-minute lecture-based session; another cohort participated in a 90-minute TBL session. Academic performance was assessed using validated pre- and post-tests. Student satisfaction and self-efficacy were evaluated using post-session surveys derived from the Kirkpatrick model and Bandura’s self-efficacy theory. Independent sample t-tests compared quantitative outcomes, and qualitative responses were analyzed thematically.

**Results:** Both cohorts demonstrated improvement in chest radiograph interpretation scores, with no statistically significant differences between groups in post-test performance or score improvement (p = 0.841). Survey results indicated favorable perceptions of both instructional approaches. The TBL cohort reported significantly higher ratings for engagement and peer interaction (p = <0.001). Self-efficacy ratings were higher among TBL participants for selected confidence-related items (p=0.003, p = 0.021, p = <0.001). Qualitative responses on what contributed most to self-efficacy emphasized peer discussion in the TBL group and structured explanations in the lecture group.

**Conclusions:** TBL produced academic performance comparable to lecture-based instruction while supporting greater learner engagement and confidence. These findings support TBL as a feasible instructional approach for chest radiograph interpretation in PA education.

## Background

In modern medicine, radiology serves as a cornerstone in the diagnosis, management, and treatment of many medical conditions. However, radiology education is often underrepresented in health professions curricula despite its critical importance (1-3). The Accreditation Review Commission on Education for the Physician Assistant (ARC-PA) mandates proficiency in diagnostic imaging as a standard for program accreditation, yet there is no literature on methodology and effectiveness of radiology education for physician associate (PA) students (4).

A larger body of research on radiology education exists for undergraduate medical students and highlights significant variability and a lack of standardized teaching methods (1, 3, 5-7). A 2024 systematic review and meta-analysis aimed to identify the most effective radiology teaching methods based on student acquisition of knowledge or skill. This study revealed heterogeneity among existing approaches but found evidence supporting active-learning and e-learning strategies (3). Active learning, such as the flipped learning model, is an emerging instructional method supported by a growing body of literature (2, 8, 9). This model involves students reviewing instructional materials independently, followed by in-class activities that promote higher-order thinking and application of knowledge.

Team-Based Learning (TBL) is an active learning strategy that has shown promise in various educational contexts, including medical education (10, 11). Defined as a structured form of small group learning, TBL facilitates student engagement through a series of activities that include individual preparation, readiness assurance, group work, and immediate feedback (10). The TBL framework emphasizes the application of knowledge to solve problems, making it particularly suitable for topics requiring higher cognitive complexity based on Bloom’s Taxonomy that are foundational for radiology interpretation (8, 12). Furthermore, studies have demonstrated that TBL, rooted in social constructivism, can enhance student accountability and foster collaborative learning environments (10, 13).

Despite the growing support for TBL, literature on its implementation in medical education, particularly in radiology, remains limited. The traditional lecture-based approach continues to dominate, often leading to passive learning experiences that may not fully equip students with the necessary interpretative skills for clinical practice (11, 14). Given the critical role of diagnostic imaging in clinical decision-making and its increasing availability, there is a clear need to explore alternative instructional methods that can better prepare PA students for real-world applications.

This study aims to evaluate the effectiveness of a TBL model compared to traditional lecture-based instruction in teaching chest radiograph interpretation to PA students during their didactic training. The primary objective is to compare students’ performance in chest radiography interpretation after TBL versus lecture-based instruction. Secondary objectives include students’ satisfaction and engagement, as well as their perceived self-efficacy.

By addressing these gaps in the literature and evaluating the effectiveness of TBL in PA education, this study aims to inform future curricular developments and enhance the training of PA students in diagnostic imaging.

## Methods

A mixed-methods, pre- and post-quasi-experimental design was employed to evaluate measurable learning outcomes, students’ perspectives on teaching methods, and their sense of self-efficacy. The study took place at Wake Forest University School of Medicine within the didactic curriculum of the PA program.

### Participants

PA students enrolled in the didactic year across two consecutive cohorts were evaluated. The 2024 cohort (control group) received lecture-based instruction, while the 2025 cohort (intervention group) participated in TBL. A convenience sampling approach was used. To assess baseline comparability, admissions data from each cohort were compared including undergraduate GPA, GRE scores, and total patient care hours before matriculation.

A priori power analysis was conducted to determine minimum detectable effect size for the primary objective. The planned comparison was mean change in test scores between the lecture and TBL cohorts, analyzed with two-sided independent-samples *t* test. Assuming a minimum of 60 students per cohort, the study had 80% power to detect a medium effect size (Cohen’s *d* =0.5).

### Ethical Considerations

The Advocate Enterprise Institutional Review Board reviewed this project and determined it to be exempt. Participation was voluntary, and no course grades were affected by participation or performance on study instruments. All responses were deidentified.

### Educational Interventions

#### Lecture-Based Instruction (Control)

Students in the lecture group attended a synchronous 90-minute instructor-led session teleconferenced across two campuses in a traditional lecture hall. There were no pre-class preparation activities. Content was delivered through a Microsoft 365 PowerPoint presentation with case examples and chest radiograph images. The instructor provided real-time explanations, and students could ask questions during or after the session. There were no formal team-based activities.

#### Team-Based Learning (Intervention)

Students in the TBL group participated in a structured 90-minute session based on the seven core TBL elements, held in a classroom with grouped tables to encourage interaction (10). Prior to class, students independently reviewed three short, recorded lectures (10 minutes each) and were given a list of resources to investigate common chest radiograph findings. During the synchronous instructional session, students completed an individual readiness assurance test (iRAT) followed by a team readiness assurance test (tRAT) in randomly assigned teams of 4–5 students. Teams received immediate feedback, and the instructor facilitated clarification discussions.

Application exercises required teams to analyze and interpret normal and abnormal chest radiographs. Following the “4S” framework (significant problem, same problem, specific choice, simultaneous reporting), teams selected answers using whiteboards and reported them simultaneously (10). The instructor provided feedback, and inter-team discussion was encouraged. A schematic of the session design is shown in **Figure 1**. All components of this session were delivered using the Intedashboard platform v3 (Queenstown, Singapore). Peer evaluation was omitted given the single session format.

**Figure 1.**
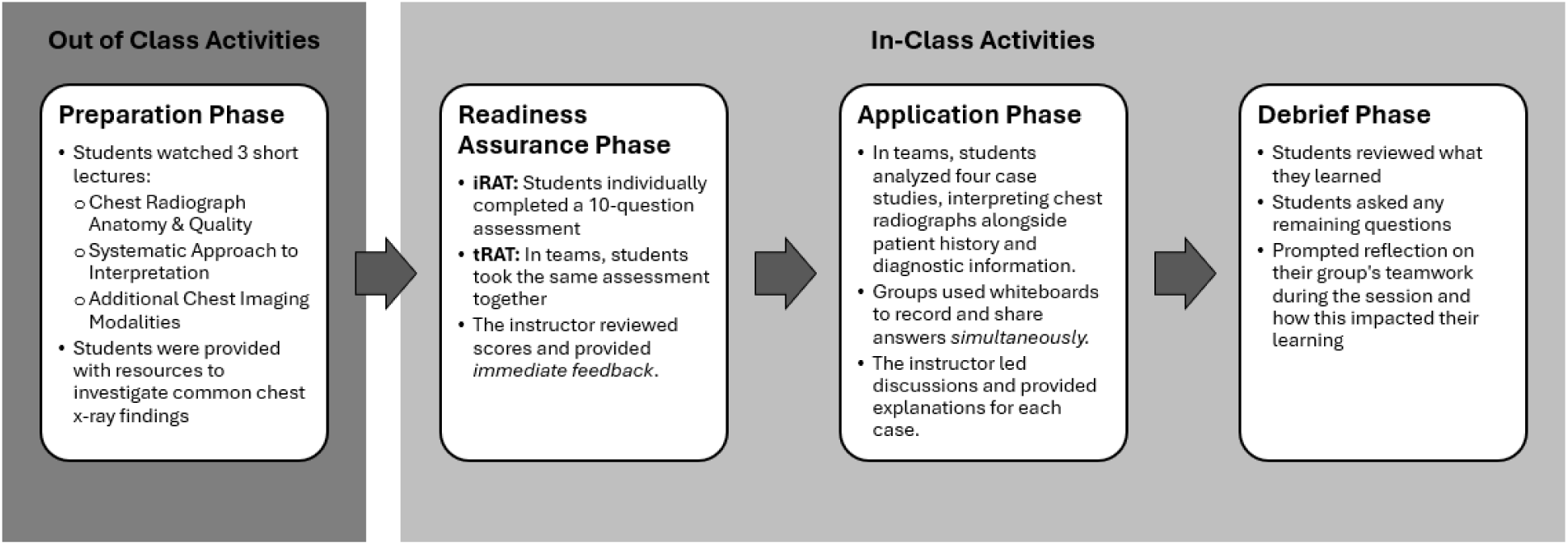
Overview of Team-Based Learning Session Structure. A visual summary of the four phases of the TBL session, Preparation, Readiness Assurance, Application, and Debrief, highlighting how students progressed from pre-class learning to collaborative case analysis and reflective discussion.

Both groups received instruction aligned to identical learning objectives, total in-class instruction time was 90 minutes and lecture-based and TBL sessions were delivered by the same instructor.

### Data Collection

#### Academic Performance

Students completed pre- and post-tests consisting of validated multiple-choice questions from the American College of Radiology (ACR) and the Alliance of Medical Student Educators in Radiology (AMSER) question bank (15). Questions were selected to match session learning objectives and administered using ExamSoft v3.10.0 (Oakland, California) online testing platform.

#### Satisfaction and Self-Efficacy

Post course survey administered via REDCap v15.5.15 (Nashville, TN) immediately following each session. Quantitative items used a 5-point Likert scale (1 = strongly disagree, 5 = strongly agree). Items on satisfaction were adapted from two established surveys the Student Evaluation of Educational Quality (SEEQ) questionnaire and the Satisfaction with Simulation Experience scale (SSES) (16, 17). Items on self-efficacy were developed based on Bandura’s Self-Efficacy Theory and the Kirkpatrick Model’s reaction and behavior levels (18, 19). Open-ended survey items elicited feedback regarding what contributed most to students’ self-efficacy in chest radiograph interpretation.

### Data Analysis

All quantitative data were analyzed using GraphPad Prism v10.6.1 (La Jolla, CA). Descriptive statistics summarized participant demographics and admissions data. Independent sample t-tests compared admissions variables across cohorts to detect potential confounders and examine baseline comparability. Descriptive statistics were used to characterize group performance and independent sample t-tests compared outcomes between groups. Survey responses were summarized using descriptive statistics, and mean Likert scores were compared using t-tests. Qualitative responses were analyzed thematically. Two independent investigators coded responses inductively, compared themes, and resolved discrepancies by consensus to ensure rigor. Analyst triangulation enhanced reliability of the findings.

## Results

### Study Population

A total of 177 PA students were invited to participate across two consecutive cohorts: 90 students in the lecture-based instruction group and 87 students in TBL group. Demographic and admissions data for each cohort are summarized in **Table 1**. There were no statistically significant differences between cohorts in age, GPA, GRE scores, or prior clinical experience hours (*p* > 0.05). Within the lecture group there was a 93% response rate for the pre- and post-tests and a 72% response rate for the survey. The TBL group had an 86% response rate for the pre- and post-tests and a 90% response rate for the survey.

**TABLE 1.** Comparison of Study Cohort Demographics and Admissions Data.

| **Variable**<br>*Mean (SD)* | **TBL**<br>*n=87* | **Lecture**<br>*n=90* | **P value** |
| --- | --- | --- | --- |
| **Age at Entry** | 25.68 (4.05) | 25.61 (3.03) | 0.901 |
| **GPA** | 3.64 (0.26) | 3.60 (0.25) | 0.336 |
| **Science GPA** | 3.51 (0.32) | 3.43 (0.36) | 0.113 |
| **Quantitative GRE Score** | 153.46 (4.84) | 152.0 (16.54) | 0.422 |
| **Verbal GRE Score** | 155.55 (5.19) | 159.90 (32.95) | 0.339 |
| **Writing GRE Score** | 4.29 (0.60) | 4.42(0.58) | 0.130 |
| **Clinical hours prior to Matriculation** | 3637 (3940) | 4142 (4197) | 0.411 |

SD = Standard deviation, GPA = grade point average, GRE = Graduate Record Examinations $p < 0.05$

### Academic Performance

Mean performance for pre- and post-tests for chest radiograph interpretation is shown in **Table 2**. Both cohorts demonstrated notable improvement from pre- to post-testing. No statistically significant between-group differences were observed in post-test scores or overall mean improvement (*p* = 0.841). The distribution of pre- and post-test scores is graphically represented in **Figure 2**.

**Figure 2.**
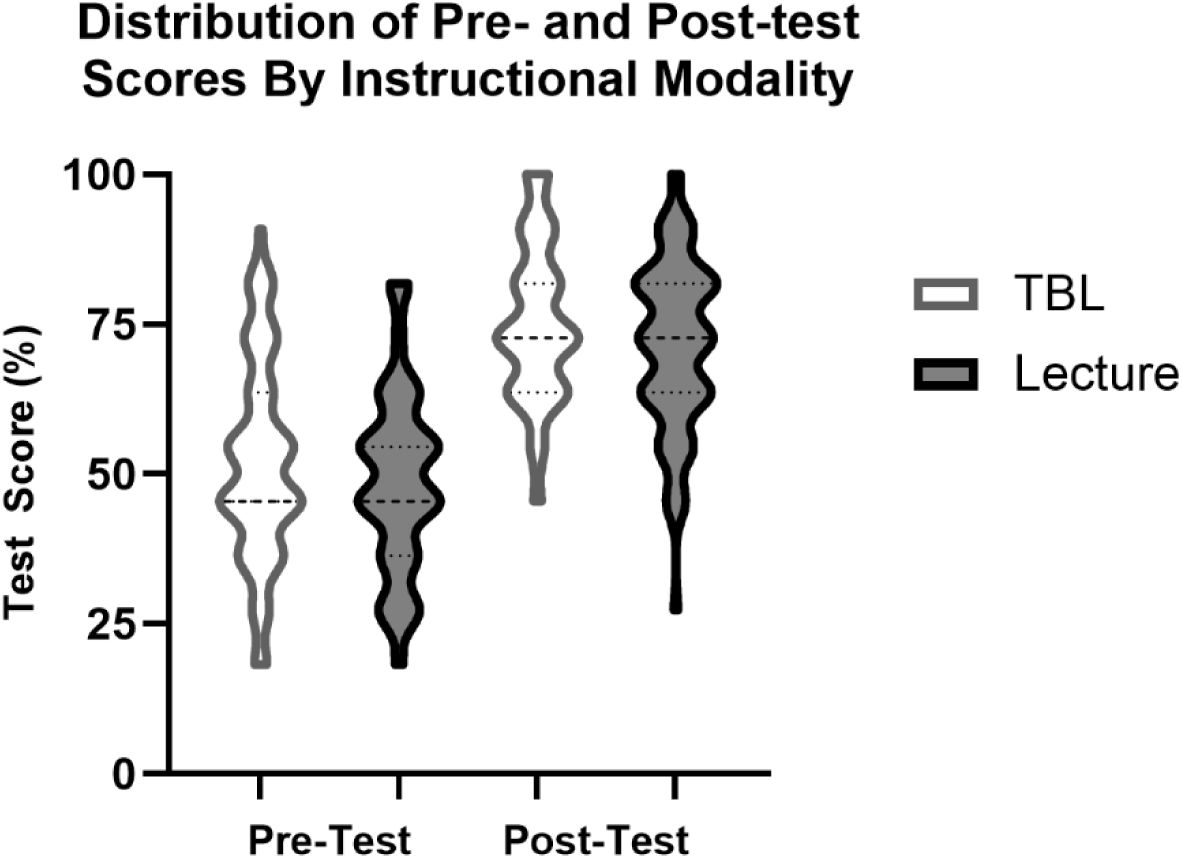
Distribution of Pre- and Post-Test Scores by Instructional Modality. Violin plots illustrate the distribution of individual student scores on the chest radiograph interpretation pre- and post-tests for both instructional modalities. The width of each violin represents score density, and the dotted lines indicate the median and interquartile range. Both cohorts demonstrated improved post-test performance, with no statistically significant difference in mean score improvement between lecture and TBL groups (*p* = 0.841).

**TABLE 2.** Comparison of Student Performance on Chest Radiology Pre- and Post-tests.

| Variable | TBL | Lecture | P Value |
| --- | --- | --- | --- |
| Mean (SD) | n=75 | n=84 |  |
| **Pre-test** | 51.88 (16.69) | 48.81 (14.92) | 0.226 |
| **Post-test** | 75.15 (13.06) | 71.54 (14.03) | 0.094 |
| **Mean Difference** | 23.27 (16.11) | 22.73 (18.10) | 0.841 |

SD = Standard deviation, TBL = Team-based Learning $p < 0.05$

### Student Satisfaction

Quantitative survey responses are summarized in **Table 3**. Both cohorts rated the sessions favorably across all satisfaction items, with mean scores exceeding 4.0 on the 5-point Likert scale. The TBL cohort reported higher mean ratings for all survey items, with statistically significant increases in five of the seven metrics assessed. Students rated overall improvement in understanding chest radiographs and the sessions’ ability to meet learning objectives similarly across both instructional modalities. The largest between-group differences were observed in engagement (*p* < 0.001) and peer/instructor interaction (*p* < 0.001), favoring TBL.

**TABLE 3.** Self-Reported Student Survey on Session Satisfaction and Self-Efficacy in Chest Radiograph Interpretation.

|  | Survey Item | TBL Mean Scores (SD)<br>n=78 | Lecture Mean Scores (SD)<br>n=65 | P Value |
| --- | --- | --- | --- | --- |
| Satisfaction | The session improved my understanding of chest radiograph interpretation. | 4.62 (0.52) | 4.48 (0.53) | 0.119 |
|  | The content of this session met the proposed learning objectives. | 4.64 (0.51) | 4.58 (0.50) | 0.505 |
|  | I was actively engaged throughout the session. | 4.72 (0.45) | 4.35 (0.67) | **<0.001** |
|  | The instructional method promoted interaction with my peers/instructor. | 4.86 (0.39) | 4.18 (0.75) | **<0.001** |
|  | The instructor presented the material in a clear and understandable manner. | 4.76 (0.51) | 4.54 (0.53) | **0.015** |
|  | The instructional method was effective for teaching chest radiograph interpretation. | 4.68 (0.59) | 4.31 (0.66) | **0.001** |
|  | I am satisfied with my overall learning experience in this session. | 4.58 (0.55) | 4.23 (0.66) | **0.001** |
| Self Efficacy | I feel confident that I can accurately identify normal findings on a chest radiograph. | 3.50 (0.73) | 3.25 (0.94) | 0.078 |
|  | I am able to recognize common pathological findings on a chest radiograph (e.g., pneumonia, pneumothorax). | 3.67 (0.66) | 3.25 (0.91) | **0.003** |
|  | Observing examples of chest radiograph interpretation improved my confidence in interpreting them myself. | 4.23 (0.68) | 4.23 (0.63) | 1.000 |
|  | Watching my peers or instructors interpret chest radiographs has enhanced my understanding. | 4.36 (0.66) | 4.08 (0.76) | **0.021** |
|  | The instructor's encouragement has increased my confidence in interpreting chest radiographs. | 4.33 (0.68) | 4.18 (0.68) | 0.195 |
|  | My peers' constructive feedback on my interpretations helped me feel more confident. | 4.17 (0.71) | 3.54 (0.81) | **<0.001** |
|  | I believe I can improve further with continued practice and support from instructors. | 4.55 (0.57) | 4.66 (0.57) | 0.278 |
|  | I have gained new skills and knowledge on chest radiograph interpretation during this session. | 4.47 (0.62) | 4.42 (0.53) | 0.539 |
|  | I understand the key principles of chest radiograph interpretation more clearly after today's session. | 4.24 (0.59) | 4.42 (0.58) | 0.082 |
|  | The teaching style in this session has enhanced my ability to retain and apply the skills I've learned, and I feel confident I will be able to use them in future clinical practice. | 4.40 (0.69) | 4.31 (0.64) | 0.420 |

Response Categories for Items Ranged From 1 (Strongly Disagree), 2 (Disagree), 3 (Neutral), 4 (Agree), to 5 (Strongly Agree). Bolded values are statistically significant, $p < 0.05$

### Self-Efficacy in Chest Radiograph Interpretation

Self-efficacy outcomes are summarized in **Table 3**. Following instruction, students in both the lecture-based and TBL cohorts reported an increase in confidence and self-efficacy in chest radiograph interpretation. TBL participants demonstrated higher average scores on eight of ten survey items measuring self-efficacy. TBL participants reported statistically significant increases on each of the following survey items: “I am able to recognize common pathological findings on a chest radiograph” (*p* = 0.003), “Watching my peers or instructors interpret chest radiographs has enhanced my understanding” (*p* = 0.021), and “My peers’ constructive feedback on my interpretations helped me feel more confident” (*p* < 0.001). Both cohorts scored similarly on all other self-efficacy metrics.

Students from both groups described elements they perceived as contributing most to their self-efficacy in chest radiograph interpretation. **Table 4** presents a summary of the thematic analysis of open-ended survey responses with illustrative quotes from each cohort. Responses from the lecture-based cohort most frequently referenced *practice cases/examples* and a *systematic approach to interpretation*. In contrast, TBL participants emphasized *peer discussion* and active engagement with *practice cases*.

**TABLE 4.** Thematic Analysis of Student Insights on Building Self-Efficacy in Chest Radiograph Interpretation.

| Theme | Relative Frequency in TBL (%)<br>n=78 | Relative Frequency in Lecture (%)<br>n=64 | TBL Group Exemplar Quotes | Lecture Group Exemplar Quotes |
| --- | --- | --- | --- | --- |
| Practice cases/examples | 32.1% | 60.0% | "Having multiple examples was really helpful to see different disease states." | "The practice cases were super helpful in improving self efficacy." |
| Systematic approach to interpretation | 7.6% | 16.9% | "Using the ABCDEFGH method was helpful to make sure I wasn't anchoring on any findings that immediately caught my eye during the x-ray interpretation!" | "Learning a stepwise process for interpreting chest radiographs." |
| Teaching method and Structure | 2.5% | 20% | "Active learning is the best way I believe that I learn! I feel confident doing this compared to just lectures." | "The session was very organized and to the point." |
| Connecting radiograph findings to pathophysiology & clinical presentation | 0% | 14.6% | _____ | "Relating clinical findings to the chest x ray images to make a diagnosis." |
| Listening to the instructor interpret images | 12.8% | 4.6% | "Having group discussion followed by the professor explaining the interpretation." | "Hearing the professor talk through each image was very helpful." |
| Discussing with peers | 62.8% | 6.2% | "Collaborating both with my classmates and being able to ask questions in real time allowed me to better identify learning gaps and fill them." | "I thought it was beneficial to hear my peers opinions and questions while navigating the various examples." |
| Reviewing material prior to class / prework | 9.0% | 3.1% | "Coming prepared from doing the prework helped me!" | "Pre-reading the lecture PowerPoint and filling out objectives prior to lecture to have a preview." |
| Readiness assessments and immediate feedback | 3.8% | 0% | "The individual readiness assessment and cases reinforced my confidence in understanding." | _____ |

## Discussion

This study explored whether TBL could serve as an effective instructional approach for teaching chest radiograph interpretation to PA students. TBL and lecture-based instruction resulted in comparable test scores, demonstrating that TBL is noninferior to lecture when measuring objective academic performance. However, TBL offered advantages in terms of student engagement, peer interaction, and increased learner confidence. These results contribute to the growing body of literature suggesting that active learning approaches can enrich radiology education without compromising academic performance or increasing faculty-to-student ratios.

Equivalent performance between TBL and lecture aligns with literature suggesting active learning yields equivalent exam outcomes while enhancing the learning experience by supporting the use of higher-order learning. In health professions education research, flipped and active learning models have shown neutral effects on exam scores, but meaningful gains in engagement, confidence, and preparedness for clinical reasoning (20, 21). In radiology instruction specifically, studies suggest that active learning fosters deeper processing of images when compared to traditional lecture-based instruction (2, 3).

The survey results highlight an important dimension beyond test scores. Students in the TBL cohort reported significantly higher engagement, interaction, and confidence in radiographic interpretation. These outcomes are essential for radiology education, as image interpretation depends on factual knowledge and confidence in using a systematic approach. Bandura’s theory of self-efficacy suggests that confidence influences motivation and performance in future tasks, especially in complex or ambiguous situations, which directly reflects the nature of clinical practice (18).

Prior studies have linked active learning with improved confidence in radiologic interpretation (11, 14). Our results further reinforce this finding. Students in the TBL cohort emphasized the value of peer discussion and immediate feedback. These elements are central to the TBL framework and help learners develop early interpretive confidence which is fundamental for clinical practice. The enhanced self-efficacy observed in the TBL group may translate into improved diagnostic reasoning behaviors over time, representing long-term gains in clinical practice.

To date, there is a paucity of data on effective approaches to radiology education for PA students (22). This is notable given that proficiency in diagnostic imaging is not only mandated as a standard for program accreditation by ARC-PA, but also recognized as a core competency for graduates (4, 23). This study provides evidence supporting the use TBL as an effective instructional method within the PA radiology curriculum.

### Implications for PA Radiology Education

This study has practical implications for PA programs seeking to strengthen radiology instruction. The data supports TBL as a viable option for PA programs aiming to incorporate more active learning without sacrificing efficiency or performance outcomes. A 2023 systematic review on barriers to active learning identifies class size, time required to develop materials, and lack of administrative support as frequent challenges across higher education (24). Similarly, health professions educators report that many active learning strategies are resource-intensive and require substantial preparation time or larger numbers of faculty, limiting feasibility in programs with large cohorts or constrained staffing (25). TBL addresses several of these concerns. It supports small-group collaboration within a large class, does not require increased faculty-to-student ratios, and relies on structured materials that can be reused across cohorts with modest updates (10). TBL represents a feasible approach even in programs with limited staffing or curricular space.

Successful implementation, however, requires attention to the learning environment. Unlike traditional lecture halls designed for passive, forward-facing instruction, TBL depends on small-group interaction. Classrooms with movable seating or grouped tables better facilitate discussion and team accountability. While TBL does not necessitate additional faculty, it does benefit from a physical layout that promotes active participation. Utilization of software designed for TBL can also improve student interface with instructional materials and streamline session flow. Platforms that support readiness assurance testing, immediate feedback, and simultaneous reporting allow faculty to monitor team performance in real time, track individual and group responses, and efficiently collect assessment data.

Second, improvements in engagement and confidence may be particularly beneficial in radiology, where PA students have limited curricular exposure and opportunities for application. Integrating TBL for select imaging modalities could help reduce the gap between didactic education and real-life practice by promoting stronger peer learning and a more deliberate analytic process when interpreting images.

Third, the findings suggest opportunities to integrate TBL across the diagnostic imaging curriculum. For example, expanding TBL to computed tomography (CT) interpretation, musculoskeletal radiographs, or ultrasound curriculum alongside hands-on practice could provide structure for repeated practice and allow students to build self-efficacy, better preparing them for clinical practice. Additionally, embedding TBL within longitudinal imaging instruction encourages a learning culture that values preparation, problem solving, and collaboration (10).

These outcomes also align with competency-based education, which emphasizes demonstrated abilities rather than short-term knowledge acquisition. Competence develops through repeated practice, application of skills in authentic contexts, and structured opportunities for feedback. The 2024 PA competencies and milestones framework highlights that observable behaviors such as clinical reasoning, communication, and professional judgment are central indicators of readiness for practice and cannot be captured through written examinations alone (26). In this study, improvements in self-efficacy, collaborative reasoning, and systematic interpretation represent meaningful progress toward these practice-ready competencies. When viewed through a competency-based lens, these gains are as important as test performance as they reflect behaviors students will rely on during clinical practice.

### Directions for Future Research

A central question is whether TBL leads to improved long-term retention of chest radiograph interpretation skills compared to traditional lecture-based methods. While this study’s test scores provided insight into short-term learning outcomes, they may not fully reflect the impact of TBL on deeper understanding and sustained knowledge retention. Future research should focus on the longitudinal impact of TBL through all phases of PA education as well as generalization to clinical practice Additionally, the feasibility of implementing TBL at scale within PA programs should be investigated. Studying the logistical and cultural factors that influence the adoption of TBL is essential for broader integration. This includes evaluating how barriers faced by faculty, curriculum planning requirements, and student preparation behaviors affect successful adoption. Gaining a clearer understanding of these elements would streamline adoption in PA programs more broadly.

Lastly, utility of TBL across all diagnostic imaging modalities should be evaluated. Chest radiographs represent a natural starting point given widespread use and are well-suited to collaborative reasoning. However, other imaging modalities may offer equally useful opportunities for further validation.

### Limitations

Several limitations should be considered when interpreting these findings. First, this study used two consecutive cohorts rather than random assignment. Although admissions data showed no significant baseline differences, unmeasured variations in cohort characteristics may have influenced outcomes. Future studies should employ random assignment to strengthen validity. Second, this study evaluated one TBL session for each group, which limits the ability to generalize the effects of TBL across more sustained instruction. Third, this study relied primarily on short-term pre- and post-tests, which may not fully capture deeper or long-term learning. Fourth, survey data are subjective and may reflect individual variability based on prior experience and perceptions.

Finally, the study took place at a single institution with one instructor delivering all sessions, which supports internal consistency but may limit generalizability. Different instructors, class sizes, or institutional cultures may influence the effectiveness of TBL and should be considered.

## Conclusion

TBL was demonstrated to be noninferior to traditional lecture-based training in chest radiograph interpretation. In addition, TBL produced stronger gains in student engagement, collaboration, and self-efficacy. These findings suggest that TBL is both an effective and feasible instructional method for PA radiology education. When viewed through the lens of competency-based education, the advantages of TBL extend beyond test performance by supporting the development of practice-ready behaviors such as applied reasoning, confidence, and collaborative problem-solving. The feasibility of TBL, particularly its ability to function without increased faculty-to-student ratios or extensive resources, positions it as a realistic option for programs aiming to incorporate active learning at scale (10). Together, these findings support the expansion of TBL within diagnostic imaging curricula and highlight the importance of evaluating instructional methods based on their ability to cultivate both knowledge and the skills required for clinical practice. Future research should explore long-term knowledge retention, generalizability of knowledge to clinical settings, and impact of repeated TBL exposure across multiple imaging modalities.

## Data Availability

All data produced in the present study are available upon reasonable request to the corresponding author.

## List of Abbreviations

ARC-PA: Accreditation Review Commission on Education for the Physician Assistant
CT: computed tomography
GPA: grade point average
GRE: Graduate Record Examinations
iRAT: Individual readiness assessment
PA: Physician Associate
SD: standard deviation
SEEQ: Student Evaluation of Educational Quality
SSES: Satisfaction with Simulation Experience Scale
TBL: Team-based learning
tRAT: Team readiness assessment

## Declarations

### Ethics approval and consent to participate

The Advocate Enterprise Institutional Review Board reviewed this project and determined it to be exempt (protocol number 00121133). Participation was voluntary, and no course grades were affected by participation or performance on study instruments. All responses were deidentified.

### Consent for publication

Not applicable.

### Availability of data and materials

The data used in this study is accessible from the corresponding author upon reasonable request.

### Authors’ contributions

KK (Lead), KC, LE and CS conceptualized and designed the study. KK collected the data, and KK and NS performed data analysis. KK drafted the manuscript. KC, LE and CS critically reviewed the manuscript. NS supervised the study and critically reviewed the manuscript.

## Acknowledgements

A pre-press copy of this manuscript has been posted to the medRxiv.org pre-print server. This manuscript was created as part of graduation requirements for the DMSc program of Wake Forest University School of Medicine Physician Associate Studies.

